# Blood pressure and antithrombotic in atrial fibrillation and coronary disease

**DOI:** 10.1101/2025.05.20.25328043

**Authors:** Shinichi Yamanaka, Takashi Noda, Kotaro Nochioka, Takumi Higuma, Koichi Kaikita, Masaharu Akao, Junya Ako, Tetsuya Matoba, Masato Nakamura, Katsumi Miyauchi, Nobuhisa Hagiwara, Kazuo Kimura, Kunihiko Matsui, Hisao Ogawa, Satoshi Yasuda, the AFIRE Investigators

## Abstract

**Backgrounds:** Atrial fibrillation (AF) is a risk factor for adverse cardiovascular outcomes in patients with stable coronary artery disease (CAD). The prognostic relevance of lower systolic blood pressure (SBP) and the optimal antithrombotic strategy in patients with AF and CAD remain to be clarified.

**Methods:** This post-hoc analysis of the AFIRE (Atrial Fibrillation and Ischemic Events with Rivaroxaban in Patients with Stable Coronary Artery Disease) trial stratified participants based on median SBP at baseline: >126 mmHg (High SBP group, n=1042) and ≤126 mmHg (Low SBP group, n=1093). The primary efficacy endpoint was a composite of cardiovascular events and all-cause death. The primary safety endpoint was major bleeding.

**Results:** The mean SBP was 139 mmHg and 114 mmHg in the High and Low SBP groups, respectively. In the propensity score-matched cohort (n=1684), the Low SBP group had a significantly higher incidence of the primary efficacy endpoint (hazard ratio [HR], 1.38; 95% confidence interval [CI], 1.01–1.88; p=0.039), while the primary safety endpoint was comparable between groups. In the Low SBP group, rivaroxaban monotherapy was associated with lower risks of both the primary efficacy (HR, 0.66; 95% CI, 0.41–0.86; p=0.006) and safety endpoints (HR, 0.40; 95% CI, 0.22–0.74; p=0.003) compared with combination therapy, whereas no significant differences were observed in the High SBP group.

**Conclusions:** Lower SBP was associated with increased risk of cardiovascular events and all-cause death. Rivaroxaban monotherapy demonstrated more favorable efficacy and safety outcomes compared with combination therapy, particularly among patients with lower SBP.

## INTRODUCTION

Various risk factors, including atrial fibrillation (AF) and low systolic blood pressure (SBP), contribute to worsening outcomes in patients with stable coronary artery disease (CAD). A systematic review and meta-analysis showed that AF independently increases the risk of death in patients with CAD^1^. Additionally, a composite of cardiovascular death, myocardial infarction, or stroke was higher in patients with stable CAD whose SBP is under 120 mmHg^2^. However, the impact of SBP in patients with both stable CAD and AF remains poorly understood.

Oral anticoagulation is essential for patients with AF due to the risk of thromboembolic events^3–6^, while antiplatelet agents are considered the cornerstone of treatment for patients with stable CAD^7–9^. As a result, combination antithrombotic therapy has frequently been used in clinical practice for patients with CAD and AF, increasing the risk of fatal and nonfatal bleeding. The AFIRE trial provided strong evidence that rivaroxaban monotherapy was noninferior to combination therapy with rivaroxaban plus a single antiplatelet agent for major adverse cardiovascular and cerebral events and superior for major bleeding in patients with AF and stable CAD, more than 1 year after revascularization (prior percutaneous coronary intervention [PCI] or coronary artery bypass grafting [CABG]) or in those with angiographically confirmed CAD not requiring revascularization^10^. However, the influence of SBP on the choice of antithrombotic therapy in this population remains unclear.

In this post-hoc analysis of the AFIRE trial, we examined whether SBP influences clinical outcomes and the choice of antithrombotic therapy in patients with AF and CAD.

## METHODS

### Study population and trial design

The AFIRE trial was a multicenter, randomized, open-label, parallel-group study. The trial design and primary results have been reported previously. ¹¹ In brief, men and women aged ≥20 years with AF and a CHADS₂ score ≥1, as well as stable coronary artery disease CAD, were enrolled. Eligible patients met one of the following criteria: (i) percutaneous coronary intervention (PCI) with or without stenting performed ≥1 year before enrollment; (ii) angiographic or coronary CT evidence of ≥50% coronary stenosis; or (iii) history of coronary artery bypass grafting (CABG) ≥1 year before enrollment. Key exclusion criteria included prior stent thrombosis, active malignancy, and poorly controlled hypertension, defined as clinic systolic blood pressure (SBP) ≥160 mmHg on two or more occasions. The trial was designed and conducted by an executive steering committee. ¹¹

The study complied with the Declaration of Helsinki and was approved by the institutional review board of the National Cerebral and Cardiovascular Center, Japan, along with the institutional review boards of all participating centers. An independent data and safety monitoring committee was responsible for study monitoring (ID: 20221366). All participants provided written informed consent. A contract research organization (Mebix) assisted with site management, data collection, statistical analysis, and manuscript preparation under the authors’ supervision.

A total of 2,240 patients were enrolled, and 2,236 were randomly assigned in a 1:1 ratio to receive rivaroxaban monotherapy (10 mg once daily for creatinine clearance 15–49 mL/min, or 15 mg once daily for creatinine clearance ≥50 mL/min) or combination therapy with rivaroxaban plus a single antiplatelet agent (aspirin or a P2Y₁₂ inhibitor), based on the treating physician’s discretion.

In this post-hoc analysis, outcomes were assessed according to systolic blood pressure (SBP) at trial entry in the modified intention-to-treat population. Of the 2,215 enrolled patients, 80 lacked SBP data, resulting in 2,135 patients available for analysis. Participants were stratified into two groups based on the median baseline SBP: the High SBP group (SBP >126 mmHg) and the Low SBP group (SBP ≤126 mmHg). To address potential confounding by comorbidities, propensity score matching (PSM) was performed using the nearest-neighbor method with a caliper width of 0.05 to balance covariates between the groups.

### Outcomes

The primary efficacy outcome was a composite of stroke, systemic embolism, myocardial infarction, unstable angina requiring revascularization, and all-cause death. The primary safety outcome was major bleeding, defined according to the criteria of the International Society on Thrombosis and Haemostasis.

### Statistical analyses

Continuous variables are presented as mean ± standard deviation or median with interquartile range, as appropriate. Categorical variables are expressed as counts and percentages. Cumulative event rates were estimated using the Kaplan–Meier method, and incidence rates were reported as percentages per patient-year. Comparisons of outcomes between the High SBP and Low SBP groups, as well as between rivaroxaban monotherapy and combination therapy within each SBP group, were conducted using Cox proportional hazards models. Results are expressed as hazard ratios (HRs) with 95% confidence intervals (CIs).

Randomization in the AFIRE trial was stratified using a minimization method based on age, sex, history of PCI, cardiac insufficiency, heart failure, hypertension, diabetes mellitus, and stroke. All statistical analyses were performed using RStudio version 2024.04.2+764 for macOS (SAS Institute, Cary, NC, USA). All reported P-values were two-sided, with values <0.05 considered statistically significant.

## RESULTS

### Baseline characteristics of the full cohort

Among 2135 patients, 1,093 were classified into Low SBP group (SBP≤126mmHg), while 1,042 patients were categorized into High SBP group (SBP>126mmHg) (**Figure 1**). **Table 1** shows the patients’ baseline characteristics. Mean SBP was 114.0 ±9.4 mmHg in Low SBP group while 139.1±10.2 mmHg in High SBP group. At baseline, the characteristics of two groups were similar, but creatinine clearance and prevalence of prior stroke were significantly lower in Low SBP group, while prior heart failure and myocardial infarction were significantly higher in Low SBP group.

**Figure 1:**
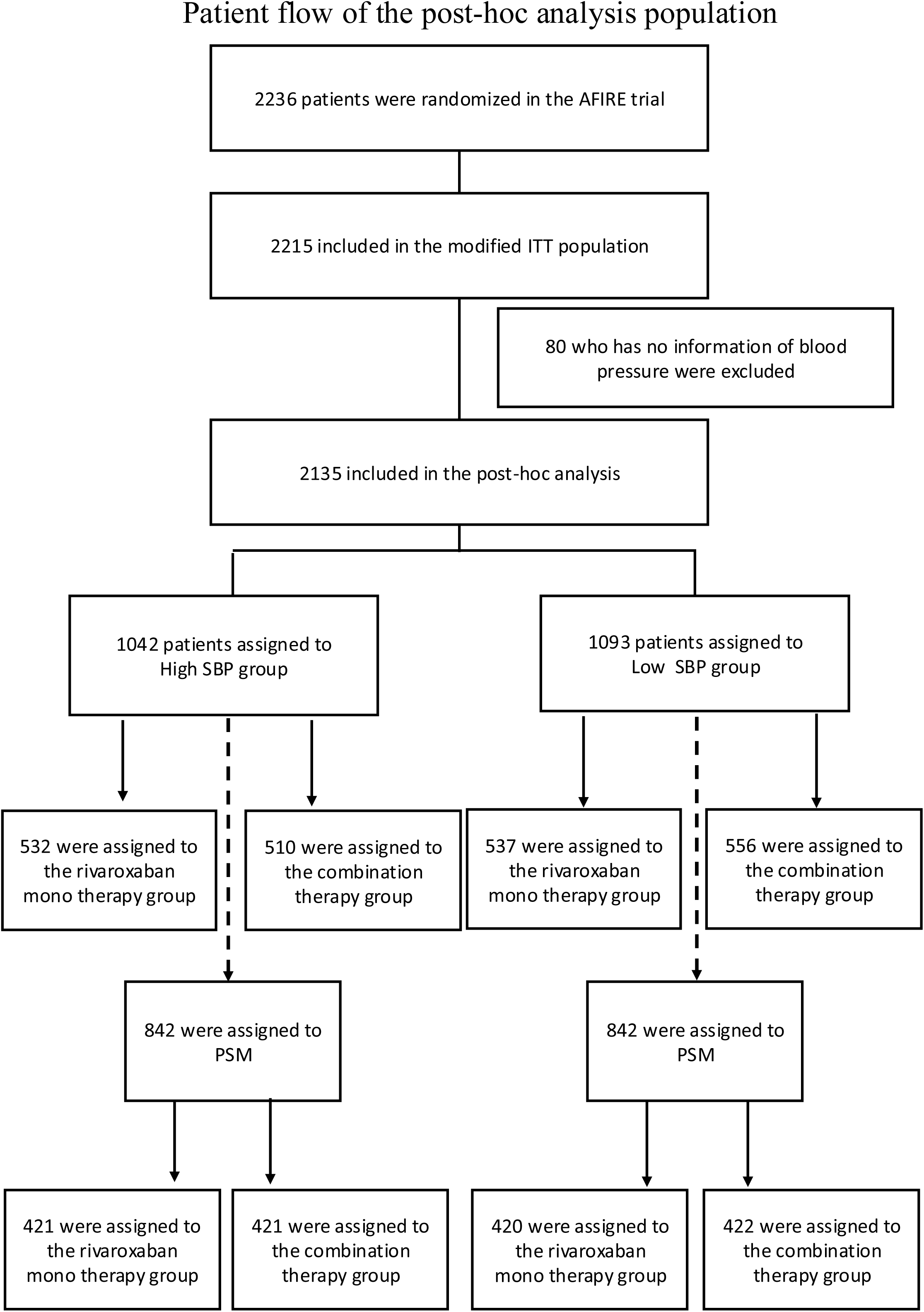
Patient flow of the post-hoc analysis population.

**Table 1:**
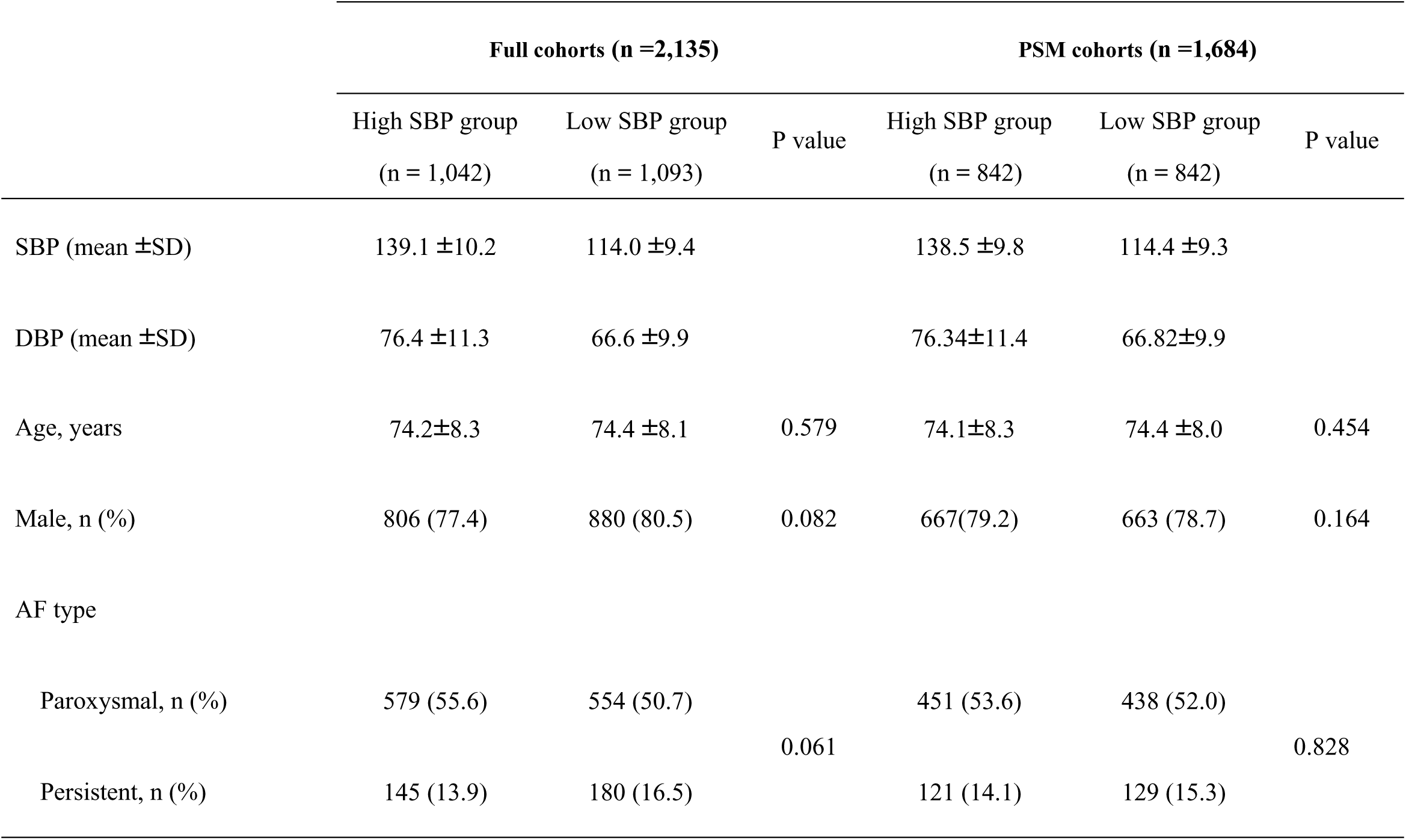
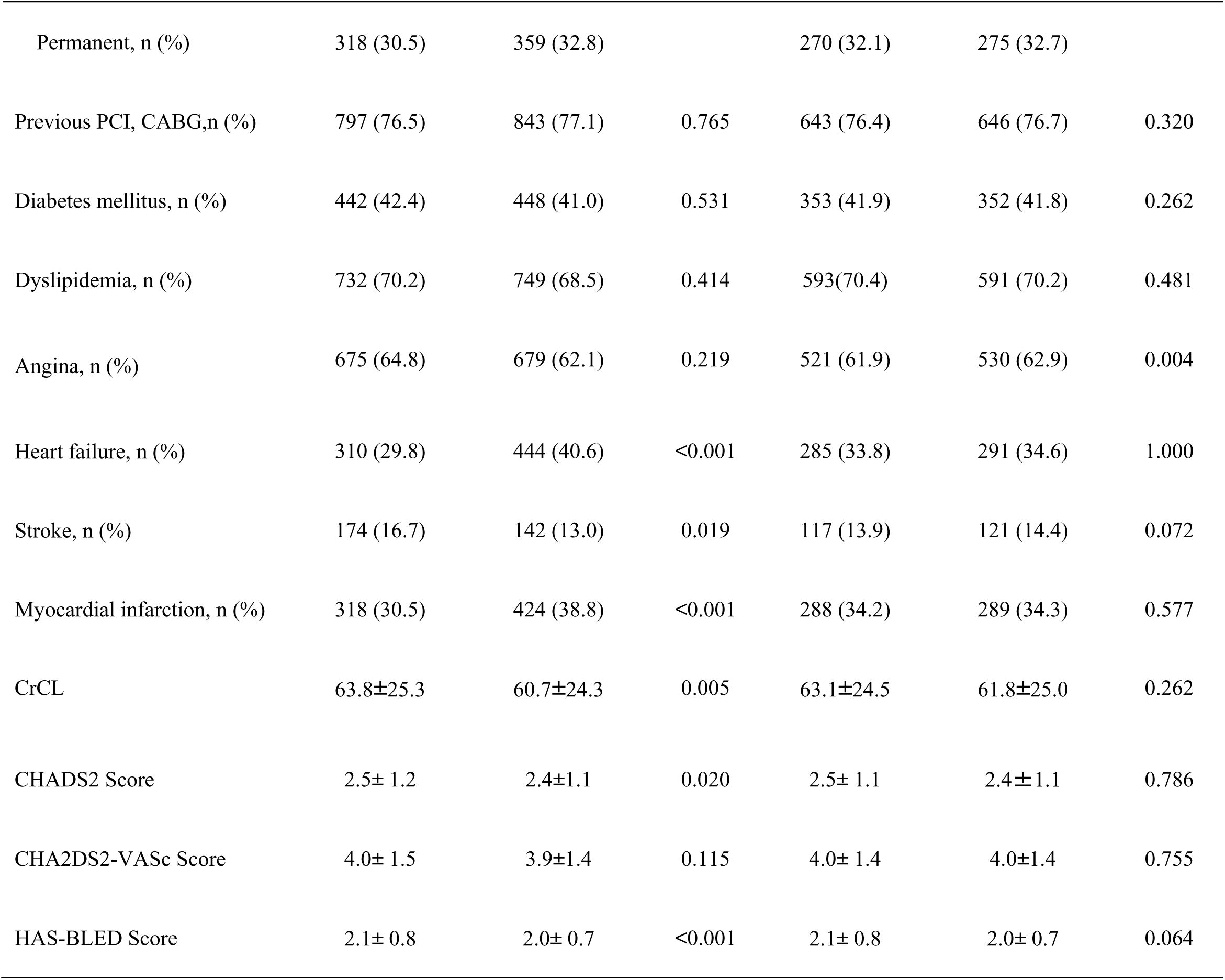
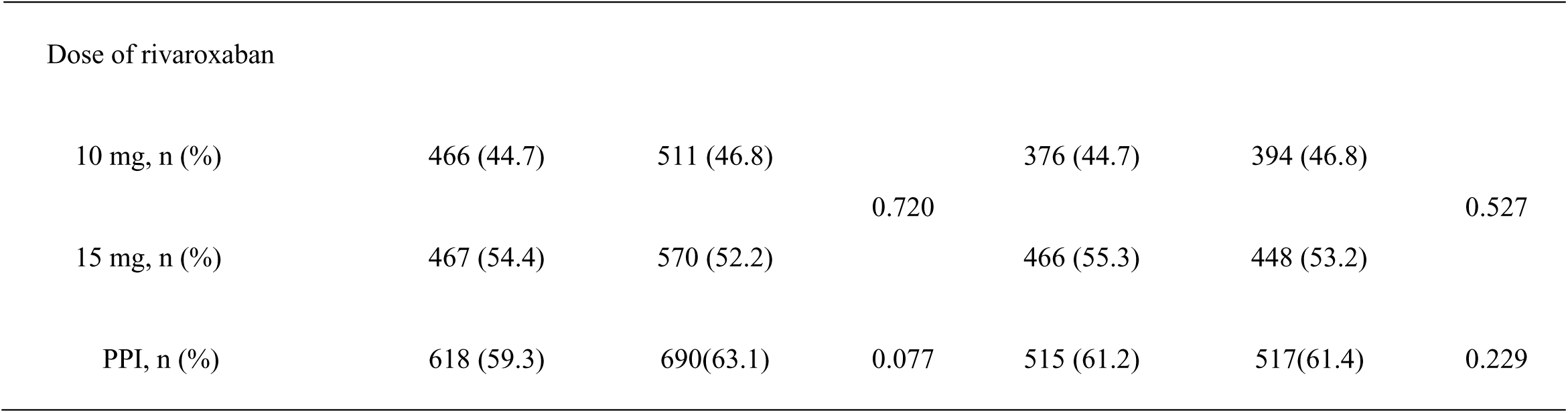
Patients’ clinical characteristics by full cohorts and propensity score-matching cohorts ‘High SBP group’ vs. ‘Low SBP group’.

3 'High SBP group' vs. 'Low SBP group'
|  | Full cohorts (n =2,135) |  |  | PSM cohorts (n =1,684) |  |  |
| --- | --- | --- | --- | --- | --- | --- |
|  | High SBP group<br>(n = 1,042) | Low SBP group<br>(n = 1,093) | P value | High SBP group<br>(n = 842) | Low SBP group<br>(n = 842) | P value |
| SBP (mean ±SD) | 139.1 ±10.2 | 114.0 ±9.4 | | 138.5 ±9.8 | 114.4 ±9.3 | |
| DBP (mean ±SD) | 76.4 ±11.3 | 66.6 ±9.9 | | 76.34±11.4 | 66.82±9.9 | |
| Age, years | 74.2±8.3 | 74.4 ±8.1 | 0.579 | 74.1±8.3 | 74.4 ±8.0 | 0.454 |
| Male, n (%) | 806 (77.4) | 880 (80.5) | 0.082 | 667(79.2) | 663 (78.7) | 0.164 |
| AF type |  |  |  |  |  |  |
| Paroxysmal, n (%) | 579 (55.6) | 554 (50.7) | 0.061 | 451 (53.6) | 438 (52.0) | 0.828 |
| Persistent, n (%) | 145 (13.9) | 180 (16.5) |  | 121 (14.1) | 129 (15.3) |  |
| Permanent, n (%) | 318 (30.5) | 359 (32.8) |  | 270 (32.1) | 275 (32.7) |  |
| Previous PCI, CABG,n (%) | 797 (76.5) | 843 (77.1) | 0.765 | 643 (76.4) | 646 (76.7) | 0.320 |
| Diabetes mellitus, n (%) | 442 (42.4) | 448 (41.0) | 0.531 | 353 (41.9) | 352 (41.8) | 0.262 |
| Dyslipidemia, n (%) | 732 (70.2) | 749 (68.5) | 0.414 | 593(70.4) | 591 (70.2) | 0.481 |
| Angina, n (%) | 675 (64.8) | 679 (62.1) | 0.219 | 521 (61.9) | 530 (62.9) | 0.004 |
| Heart failure, n (%) | 310 (29.8) | 444 (40.6) | <0.001 | 285 (33.8) | 291 (34.6) | 1.000 |
| Stroke, n (%) | 174 (16.7) | 142 (13.0) | 0.019 | 117 (13.9) | 121 (14.4) | 0.072 |
| Myocardial infarction, n (%) | 318 (30.5) | 424 (38.8) | <0.001 | 288 (34.2) | 289 (34.3) | 0.577 |
| CrCL | 63.8±25.3 | 60.7±24.3 | 0.005 | 63.1±24.5 | 61.8±25.0 | 0.262 |
| CHADS2 Score | 2.5± 1.2 | 2.4±1.1 | 0.020 | 2.5± 1.1 | 2.4±1.1 | 0.786 |
| CHA2DS2-VASc Score | 4.0± 1.5 | 3.9±1.4 | 0.115 | 4.0± 1.4 | 4.0±1.4 | 0.755 |
| HAS-BLED Score | 2.1± 0.8 | 2.0± 0.7 | <0.001 | 2.1± 0.8 | 2.0± 0.7 | 0.064 |
| Dose of rivaroxaban |  |  |  |  |  |  |
| 10 mg, n (%) | 466 (44.7) | 511 (46.8) | 0.720 | 376 (44.7) | 394 (46.8) | 0.527 |
| 15 mg, n (%) | 467 (54.4) | 570 (52.2) |  | 466 (55.3) | 448 (53.2) |  |
| PPI, n (%) | 618 (59.3) | 690(63.1) | 0.077 | 515 (61.2) | 517(61.4) | 0.229 |

### Primary efficacy and safety endpoints in the full cohort: comparison between Low SBP and High SBP groups

Figures 2A and **2B** show the primary efficacy and safety endpoints between Low SBP group and High SBP group in full cohorts. Primary efficacy events were significantly higher in Low SBP group than in High SBP group (HR, 1.50; 95% CI, 1.13–1.98; P=0.004). With respect to primary safety events, there was no statistically difference in Low SBP group compared with High SBP group (HR, 1.30; 95% CI, 0.89–2.07; P=0.14). With respect to the primary efficacy events, the events risk was higher in Low SBP group than in High SBP group across in most of the subgroups. Data are shown in **Supplementary** Figure 1. **Supplementary Table 1** shows the Individual components of events. Unstable angina pectoris and death were especially higher in Low SBP group compared to High SBP group.

**Figure 2:**
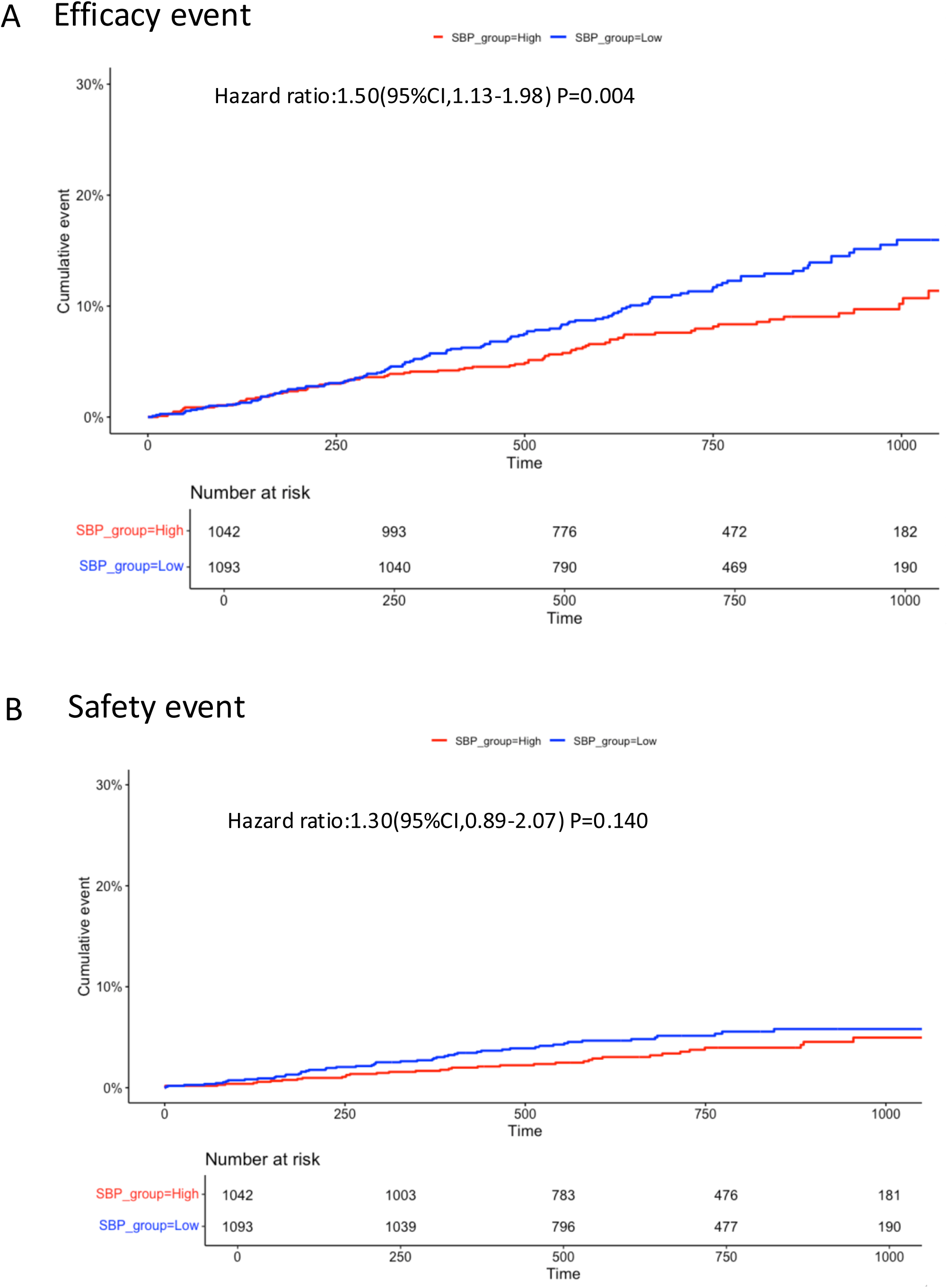
Kaplan-Meier curves for efficacy and safety events in Low SBP group and High SBP Group from the full cohorts. Panel A shows the efficacy events (composite of stroke, systemic embolism, myocardial infarction, unstable angina requiring revascularization, and death of any cause) in Low SBP group and High SBP group from the full cohort. Panel B shows the safety events for major bleeding in Low SBP group and High SBP group from the full cohort.

### Baseline characteristics of the PSM cohort

The propensity score was estimated using a logistic regression model, considering covariates such as age, sex, AF type, previous revascularization, diabetes mellitus, dyslipidemia, angina, heart failure, myocardial infarction, creatinine clearance, dose of rivaroxaban, and use of PPI. Finally, 842 patients were extracted from each group (Figure 1). The baseline characteristics of the PSM cohort are also summarized in **Table 1**. Mean SBP was 114 ±9 mmHg in Low SBP group while 139 ±10 mmHg in High SBP group. The baseline covariates were well balanced between the groups, except for a higher prevalence of a history of angina in Low SBP group.

### Primary efficacy and safety endpoints in the PMS cohort; comparison between Low SBP and High SBP groups

Figures 3A and **3B** show the primary efficacy and safety endpoints between Low SBP group and High SBP group in PSM cohort. The primary efficacy events were significantly higher in Low SBP group than in High SBP group (HR, 1.38; 95% CI, 1.01–1.88; p=0.039) and equivalent safety events (HR, 1.45; 95% CI, 0.43–1.08; p=0.140).

**Figure 3:**
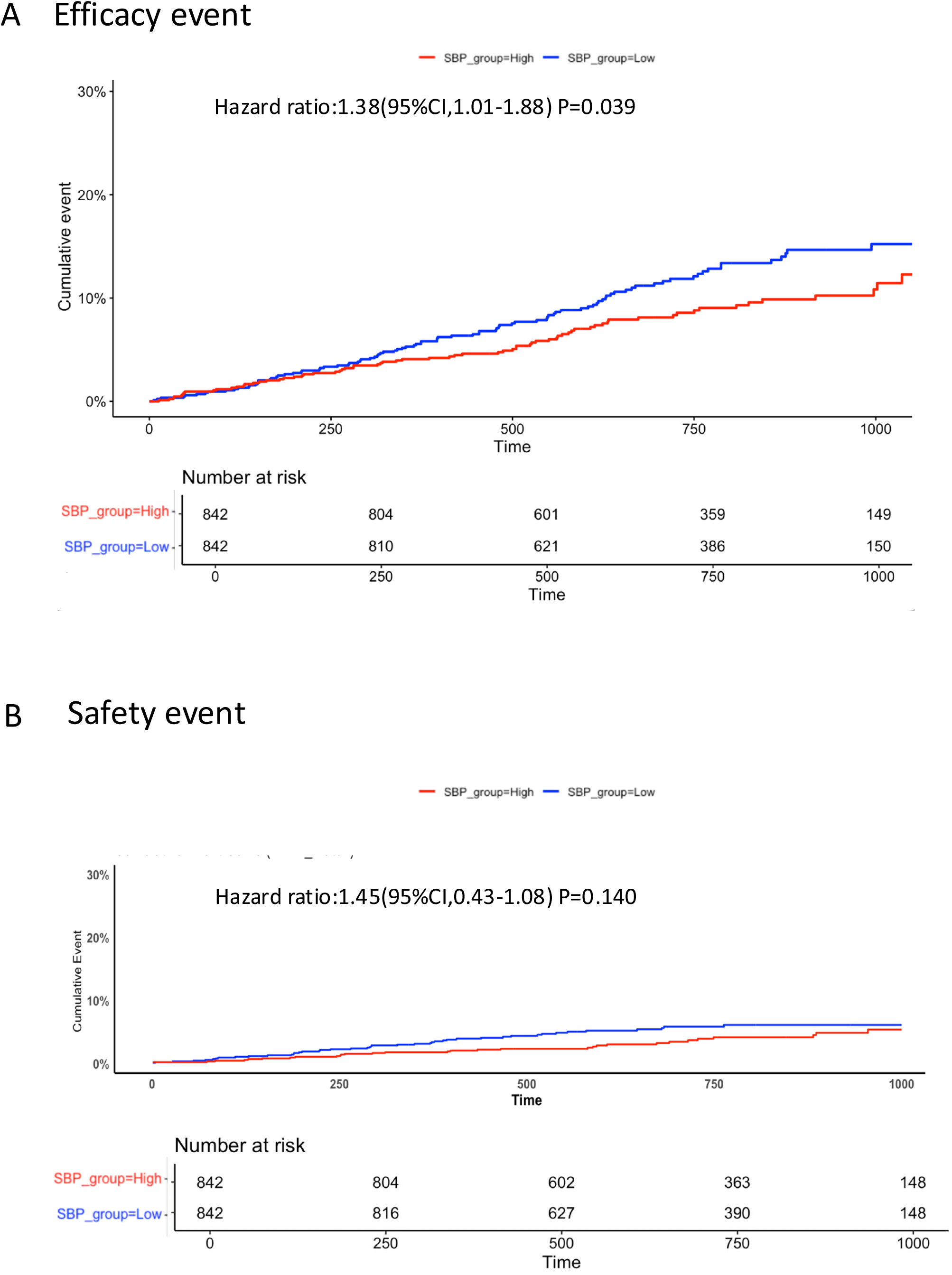
Kaplan-Meier curves for efficacy and safety events in Low SBP group and High SBP Group from the PSM cohort. Panel A shows the efficacy events (composite of stroke, systemic embolism, myocardial infarction, unstable angina requiring revascularization, and death of any cause) in Low SBP group and High SBP group from PSM cohorts. Panel B shows the safety events for major bleeding in Low SBP group and High SBP group from PSM cohorts.

### Clinical significance of monotherapy in Low SBP and High SBP groups

Figures 4A and **4B** show the difference of the primary efficacy endpoints between monotherapy and combination therapy in Low SBP group and High SBP group. Here, we used the full cohort data. In Low SBP group, monotherapy was superior to combination therapy regarding primary efficacy endpoints (HR, 0.60; 95% CI, 0.41–0.86; P=0.006, Figure 4A), while there was no statistical difference between monotherapy and combination therapy in High SBP group (Figure 4B).

**Figure 4:**
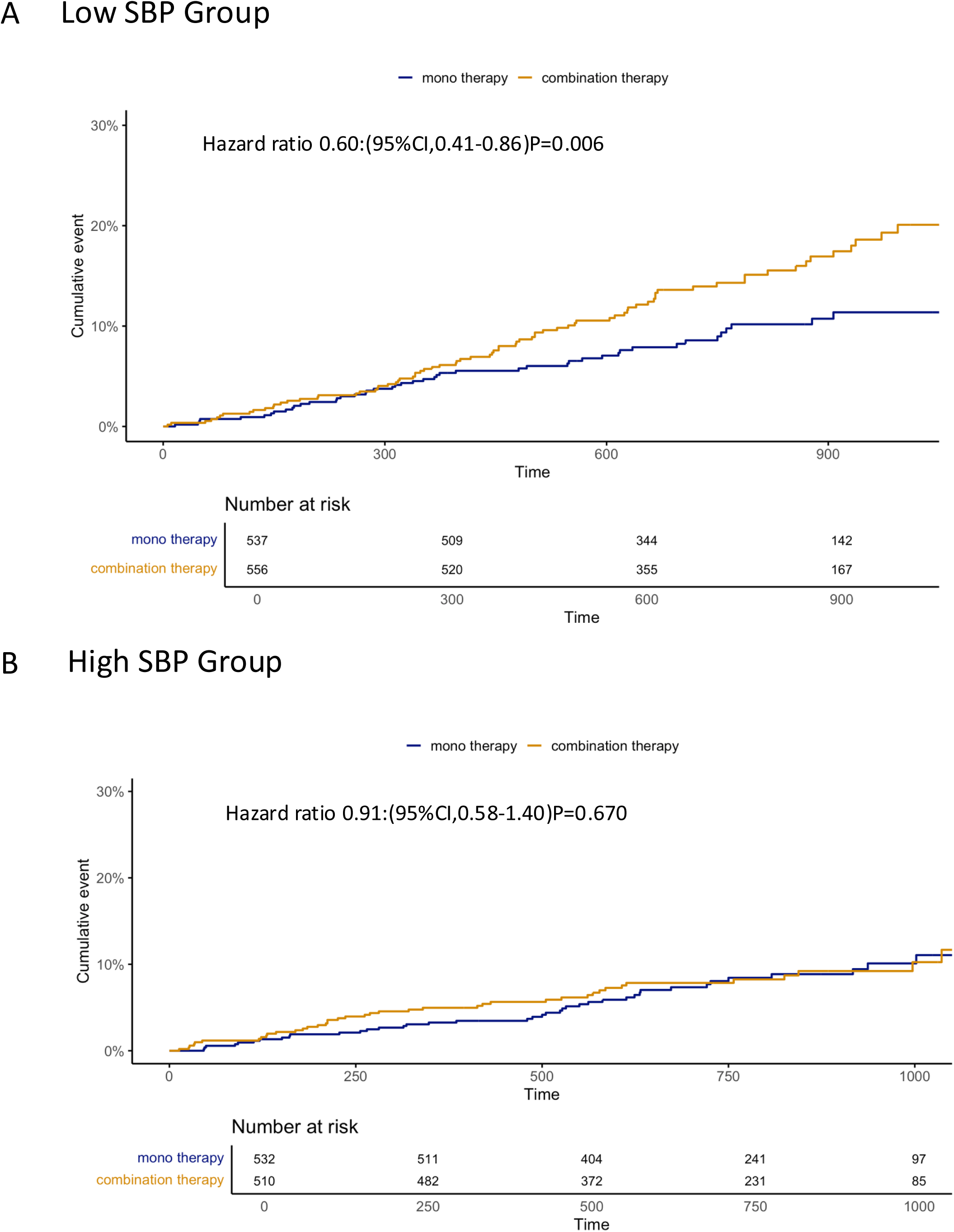
Kaplan-Meier curves for efficacy events in Low SBP group and High SBP between monotherapy and combination therapy. Panel A shows efficacy events between monotherapy and combination therapy in Low SBP group. Panel B shows efficacy events between monotherapy and combination therapy in High SBP group.

Regarding the primary safety endpoints, monotherapy was also superior to combination therapy in Low SBP grou (p HR, 0.40; 95% CI, 0.22–0.74; P=0.003, Figure 5A). However, in High SBP group, no statistical difference between monotherapy and combination therapy was also observed (HR 0.99; 95%CI, 0.52-1.89; P=0.98, Figure 5B).

**Figure 5:**
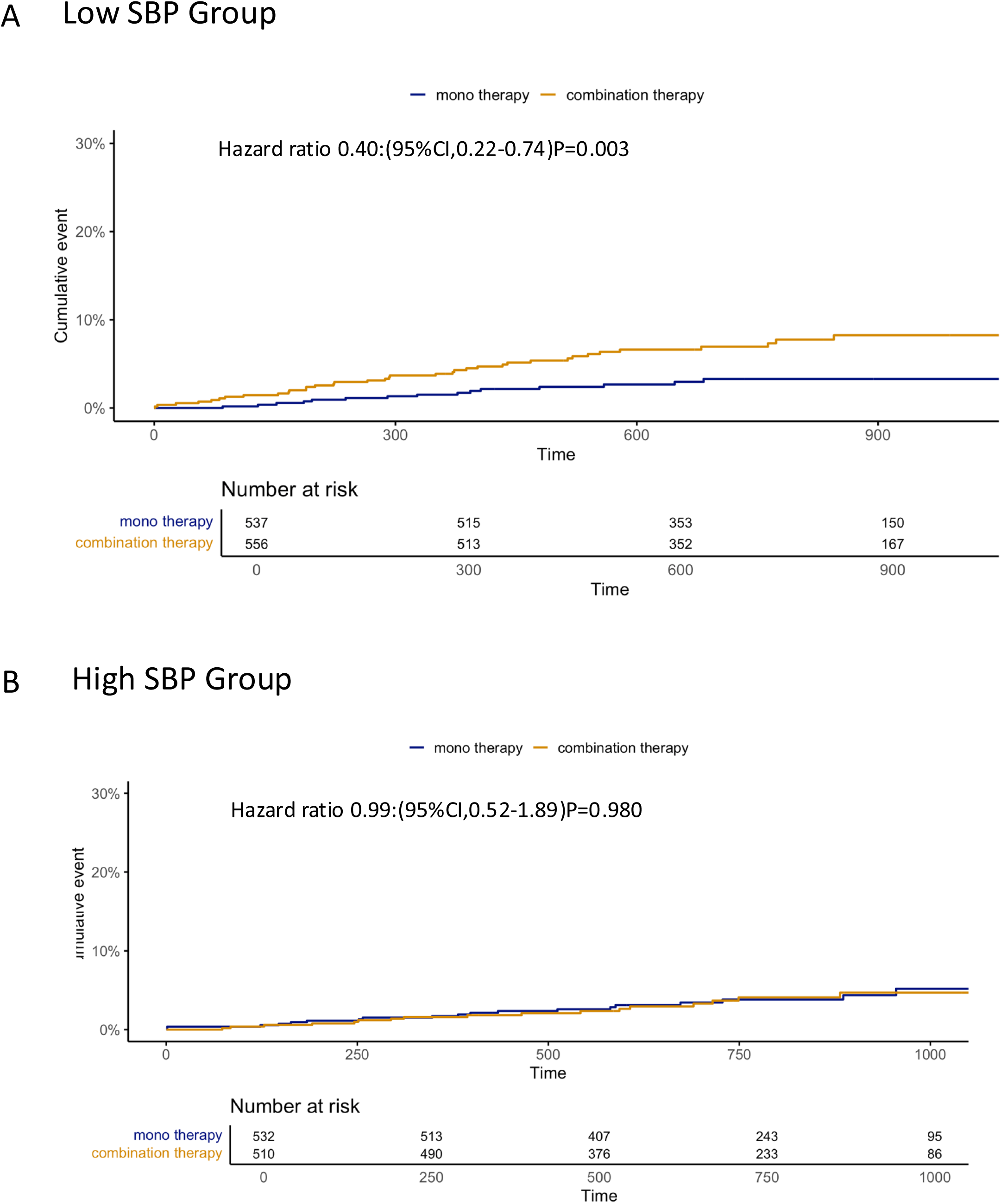
Kaplan-Meier curves for safety events in Low SBP group and High SBP between monotherapy and combination therapy. Panel A shows safety events between monotherapy and combination therapy in Low SBP group. Panel B shows safety events between monotherapy and combination therapy in High SBP group.

**Supplementary Table 2** and **3** show individual components of efficacy and safety events respectively between monotherapy and combination therapy in each SBP group. Among the Low SBP group, events of cerebral infarction and death were significantly lower in monotherapy group than in combination therapy group while individual components were similar in High SBP group.

## DISCUSSION

In this post-hoc analysis of the AFIRE trial, the major findings were as follows: 1) Low SBP was independently associated with a higher risk of the primary efficacy endpoint. 2) Rivaroxaban monotherapy was associated with favorable efficacy and safety outcomes, particularly in patients with low SBP.

### Clinical impact of SBP on primary efficacy endpoints

The optimal SBP range for patients with stable CAD and concomitant AF remains unclear. Previous studies have reported an increased risk of cardiovascular death, myocardial infarction, or stroke in patients with stable CAD when SBP is below 120 mmHg². Additionally, recent data suggest that in patients with AF, all-cause mortality is elevated when SBP is below 115 mmHg¹². Low SBP may reduce perfusion to vital organs, particularly in patients with stable CAD and AF, where compensatory cardiac function is often impaired.

In this post-hoc analysis of the AFIRE trial, the incidence of primary efficacy events was significantly higher in the low SBP group, consistent with previous studies reporting poorer clinical outcomes associated with lower blood pressure²,¹². Patients with lower SBP are more likely to exhibit diminished systemic perfusion, particularly when cardiac function is already compromised. Furthermore, the low SBP group in our cohort had a greater prevalence of reduced creatinine clearance, prior myocardial infarction, and a history of heart failure—conditions that are closely linked to increased clinical vulnerability. These factors, in combination with low SBP, may contribute to the elevated event rates observed.

To further evaluate the influence of SBP independent of baseline comorbidities, a PSM analysis was performed. Even after adjustment, the low SBP group demonstrated a significantly higher incidence of efficacy endpoints, suggesting that lower SBP itself is associated with worse outcomes in this population. This underscores the need for careful consideration of SBP levels in patients with stable CAD and AF. Maintaining an appropriate SBP range may help support adequate organ perfusion and improve overall prognosis in this complex clinical setting.

**Supplementary** Figure 2 illustrates the association between SBP and the risk of primary efficacy events in the AFIRE population. Data from the prospective observational longitudinal registry of patients with stable coronary artery disease (CLARIFY) registry demonstrated that not only low but also elevated SBP levels are associated with increased cardiovascular risk in patients with stable CAD². For instance, compared with a reference SBP group of 120–129 mmHg, the HR for the primary outcome was 1.51 (95% CI, 1.32–1.73) in patients with SBP of 140–149 mmHg, and 2.48 (95% CI, 2.14–2.87) in those with SBP ≥150 mmHg. Moreover, Systolic Blood Pressure Intervention Trial (SPRINT) demonstrated that intensive SBP lowering to <120 mmHg, compared with a target of <140 mmHg, reduced fatal and nonfatal cardiovascular events and all-cause mortality among high-risk individuals without diabetes¹³. Similar benefits of intensive blood pressure control have been reported in patients with type 2 diabetes as well¹⁴.

In contrast, the present post-hoc analysis of the AFIRE trial did not show a clear U-shaped association between SBP and the risk of primary efficacy events. Rather, the hazard ratio curve demonstrated a steady downward trend with decreasing SBP. This pattern may be attributable in part to the trial’s exclusion criteria, which ruled out patients with uncontrolled hypertension—defined as a clinic SBP ≥160 mmHg on two or more occasions—due to concerns about bleeding risk associated with antithrombotic therapy. Consequently, the number of patients with SBP ≥160 mmHg was quite small, limiting the ability to assess the potential risks associated with markedly elevated SBP in this study population.

### Clinical impact of SBP on primary safety endpoints

Regarding primary safety events, the HAS-BLED score was significantly lower in the Low SBP group compared with the High SBP group (Low SBP group: 2.0 ± 0.7, High SBP group: 2.2 ± 0.8, P < 0.001). However, there was no significant difference in the primary safety endpoints between the Low SBP and High SBP groups. In another post-hoc analysis of the AFIRE trial, no difference in HAS-BLED scores was observed between patients with bleeding events and those without bleeding events ^15^. While the HAS-BLED score was designed to assess bleeding risk in patients on warfarin, it did not effectively differentiate bleeding risk in the AFIRE trial, despite a higher prevalence of previous myocardial infarction and/or heart failure in the Low SBP group, conditions that could increase the risk of bleeding ^16, 17^.

### Clinical significance of monotherapy in Low SBP group

As bleeding is closely associated with subsequent cardiovascular events in patients with CAD ^18^, minimizing bleeding risk is essential in the context of antithrombotic therapy. A recent sub-analysis of the AFIRE trial indicated that patients who experienced major bleeding had a higher incidence of cardiovascular events, and that major bleeding was associated with increased morbidity and mortality ^15^. In the present analysis, the Low SBP group included more patients with a history of myocardial infarction and/or heart failure, factors that may contribute to an elevated bleeding risk.

In this higher-risk population, rivaroxaban monotherapy may offer a clinical advantage by reducing both efficacy and safety events compared with combination therapy. The original AFIRE trial demonstrated that rivaroxaban monotherapy was noninferior to combination therapy regarding efficacy, and superior with respect to safety, in patients with AF and stable CAD ^10^. In the current post-hoc analysis, the benefit of monotherapy appeared to be particularly pronounced in the Low SBP group, which included patients with more comorbidities. These findings support the use of rivaroxaban monotherapy in this subgroup as a reasonable strategy to balance ischemic and bleeding risks.

### Limitations

This study has several limitations. First, although the overall sample size was large, dividing the cohort into subgroups reduced the number of patients available for each analysis, which may have limited statistical power. Second, SBP was assessed only at baseline; thus, changes in SBP during the follow-up period were not captured, leaving the longitudinal effects of blood pressure uncertain. Third, patients with uncontrolled hypertension—defined as a clinic SBP ≥160 mmHg on two or more occasions—were excluded from the trial. This exclusion likely reduced the incidence of adverse events, particularly in the High SBP group, potentially influencing the comparative analysis. Fourth, the Low SBP group may have included a heterogeneous population, such as patients with well-controlled blood pressure and others with underlying clinical conditions contributing to low SBP, which could have influenced outcomes.

### Conclusion

This post-hoc analysis of the AFIRE trial demonstrated that low systolic blood pressure was independently associated with a higher risk of efficacy events in patients with AF and stable CAD. In this high-risk population, rivaroxaban monotherapy was associated with more favorable safety and efficacy outcomes than combination therapy, supporting its use in clinical practice.

## PERSPECTIVES

AF has been shown to independently increase the risk of death in patients with CAD, as demonstrated in a recent systematic review and meta-analysis. In addition, lower SBP, particularly values below 120 mmHg, has been associated with increased cardiovascular risk in patients with stable CAD. However, the prognostic implications of SBP in patients with both AF and stable CAD have not been fully elucidated. This post-hoc analysis of the AFIRE trial provides new insight into the association between baseline SBP and clinical outcomes in this population. Lower SBP was associated with a higher risk of cardiovascular events and all-cause mortality. Rivaroxaban monotherapy was associated with more favorable efficacy and safety outcomes compared with combination therapy, particularly in patients with lower SBP. These findings suggest that SBP may be an important factor to consider when selecting antithrombotic therapy in clinical practice. Nonetheless, the post-hoc nature of the analysis and limitations in the assessment of blood pressure warrant cautious interpretation. Prospective studies with standardized and precise blood pressure measurement are needed to confirm these findings and inform optimal treatment strategies in patients with AF and stable CAD.

## Data Availability

The data that support the findings of this study are available from the corresponding author upon reasonable request.

## ACKNOWLEDGEMENTS

The author thanks all the members of AFIRE investigator.

## SOURCES OF FUNDING

This work was supported by the Japan Cardiovascular Research Foundation based on a contract with Bayer Yakuhin, Ltd., which had no role in the study design, data collection or analysis, results interpretation, or manuscript writing.

Dr. Noda reports Grants-in-Aid for Scientific Research (22K08092) from the Ministry of Education, Culture, Sports, Science, and Technology of Japan and personal fees from Bayer Yakuhin, Medtronic Japan, and Biotronik Japan. Dr. Nochioka reports grants from the Japan Agency for Medical Research and Development (AMED; 21ek0210136h0003). Dr. Kaikita reports trust research/joint research funds from Bayer Yakuhin, and Daiichi-Sankyo; scholarship funds from Abbott; and personal fees from Bayer Yakuhin, Daiichi-Sankyo, Novartis Pharma AG, and Otsuka Pharmaceutical Co. Dr. Akao reports grants from the Japan AMED, personal fees from Bristol Myers Squibb and Nippon Boehringer Ingelheim, and grants and personal fees from Bayer Yakuhin and Daiichi-Sankyo. Dr. Ako reports personal fees from Bayer Yakuhin and Sanofi and grants and personal fees from Daiichi-Sankyo. Dr. Matoba reports grants from the Japan Cardiovascular Research Foundation and personal fees from Nippon Boehringer Ingelheim, Daiichi-Sankyo, AstraZeneca, and Bayer Yakuhin. Dr. Nakamura reports grants and personal fees from Bayer Yakuhin, Daiichi-Sankyo, and Sanofi and personal fees from Bristol Myers Squibb and Nippon Boehringer Ingelheim. Dr. Miyauchi reports personal fees from Amgen Astellas BioPharma, Astellas Pharma, MSD, Bayer Yakuhin, Sanofi, Takeda Pharmaceutical, Daiichi-Sankyo, Nippon Boehringer Ingelheim, and Bristol Myers Squibb. Dr. Hagiwara reports grants and personal fees from Bayer Yakuhin, grants from Nippon Boehringer Ingelheim, and personal fees from Bristol Myers Squibb. Dr. Kimura reports grants from the Japan Cardiovascular Research Foundation, grants and personal fees from Bayer Yakuhin, Daiichi-Sankyo, Sanofi, MSD, and AstraZeneca, and personal fees from Bristol Myers Squibb and Nippon Boehringer Ingelheim. Dr. Ogawa reports personal fees from Abbott Medical Japan, Bayer Yakuhin, Daiichi-Sankyo, Eisai, Kowa, Takeda Pharmaceutical, and Teijin. Dr. Yasuda reports on grants from Takeda Pharmaceutical, Abbott, and Boston Scientific and personal fees from Daiichi-Sankyo and Bristol Myers Squibb. The other authors declare no conflicts of interest for this study.

## DISCLOSURES

None.

## NOVELTY AND RELEVALANCE

### What is new?

Lower systolic blood pressure (SBP) is independently associated with worse outcomes in patients with atrial fibrillation (AF) and stable coronary artery disease (CAD). Rivaroxaban monotherapy was more effective and safer than combination therapy, especially in those with low SBP and comorbidities such as myocardial infarction, renal dysfunction, or heart failure.

### What is relevant to hypertension?

Low SBP worsens prognosis in patients with AF and stable CAD and may influence the effectiveness of antithrombotic therapy.

### Clinical/Pathophysiological implications?

In patients with low SBP, AF, and stable CAD, careful monitoring of blood pressure is important. Rivaroxaban monotherapy should be considered as a preferred treatment option to improve both efficacy and safety outcomes.

## ABBREVIATIONS AND ACRNYMS

AF: atrial fibrillation
AFIRE: Atrial Fibrillation and Ischemic Events with Rivaroxaban in Patients with Stable Coronary Artery Disease
CABG: coronary artery bypass grafting
CAD: coronary artery disease
CI: confidence interval
HR: hazard ratio
PCI: percutaneous coronary intervention
SBP: systolic blood pressure
BMI: body mass index
CrCL: creatinine clearance
PPI: proton pump inhibitor
ITT: intention-to-treat
PSM: propensity score matching

## FIGURE LEGENDS

**Supplementary Figure 1:** Subgroup analysis of efficacy events in Low SBP group and High SBP group in full cohort.

**Supplementary Figure 2:** Association between systolic blood pressure (SBP) and risk of primary efficacy events in the full cohort. Panel A illustrates the non-linear relationship between baseline SBP and the hazard ratio (HR) for primary efficacy events. Panel B displays the distribution of efficacy events across SBP categories, with black bars indicating patients with events and blue bars indicating those without.

## Notes

### Competing Interest Statement

The authors have declared no competing interest.

### Clinical Trial

The study was approved by the Institutional Review Board of the Tohoku University Graduate School of Medicine (Approval ID: 20221366), Japan.

### Funding Statement

The authors received no external funding for this work.

## REFERENCES

1. Saglietto A, Varbella V, Ballatore A, Xhakupi H, Ferrari GM, Anselmino M. Prognostic implications of atrial fibrillation in patients with stable coronary artery disease: a systematic review and meta-analysis of adjusted observational studies. Rev Cardiovasc Med. 2021;22:439–444. doi: 10.31083/j.rcm2202049

2. Vidal-Petiot E, Ford I, Greenlaw N, Ferrari R, Fox KM, Tardif JC, Tendera M, Tavazzi L, Bhatt DL, Steg PG, et al. Cardiovascular event rates and mortality according to achieved systolic and diastolic blood pressure in patients with stable coronary artery disease: an international cohort study. Lancet. 2016;388:2142–2152. doi: 10.1016/S0140-6736(16)31326-5

3. Ono K, Iwasaki YK, Akao M, Ikeda T, Ishii K, Inden Y, Kusano K, Kobayashi Y, Koretsune Y, Sasano T, et al. JCS/JHRS 2020 Guideline on Pharmacotherapy of Cardiac Arrhythmias. J Arrhythm. 2022;38:833–973. doi: 10.1002/joa3.12714

4. January CT, Wann LS, Calkins H, Chen LY, Cigarroa JE, Cleveland JC, Jr., Ellinor PT, Ezekowitz MD, Field ME, Furie KL, et al. 2019 AHA/ACC/HRS Focused Update of the 2014 AHA/ACC/HRS Guideline for the Management of Patients With Atrial Fibrillation: A Report of the American College of Cardiology/American Heart Association Task Force on Clinical Practice Guidelines and the Heart Rhythm Society. J Am Coll Cardiol. 2019;74:104–132. doi: 10.1016/j.jacc.2019.01.011

5. Hindricks G, Potpara T, Dagres N, Arbelo E, Bax JJ, Blomstrom-Lundqvist C, Boriani G, Castella M, Dan GA, Dilaveris PE, et al. 2020 ESC Guidelines for the diagnosis and management of atrial fibrillation developed in collaboration with the European Association for Cardio-Thoracic Surgery (EACTS): The Task Force for the diagnosis and management of atrial fibrillation of the European Society of Cardiology (ESC) Developed with the special contribution of the European Heart Rhythm Association (EHRA) of the ESC. Eur Heart J. 2021;42:373–498. doi: 10.1093/eurheartj/ehaa612

6. Angiolillo DJ, Bhatt DL, Cannon CP, Eikelboom JW, Gibson CM, Goodman SG, Granger CB, Holmes DR, Lopes RD, Mehran R, et al. Antithrombotic Therapy in Patients With Atrial Fibrillation Treated With Oral Anticoagulation Undergoing Percutaneous Coronary Intervention: A North American Perspective: 2021 Update. Circulation. 2021;143:583–596. doi: 10.1161/CIRCULATIONAHA.120.050438

7. Nakamura M, Kimura K, Kimura T, Ishihara M, Otsuka F, Kozuma K, Kosuge M, Shinke T, Nakagawa Y, Natsuaki M, et al. JCS 2020 Guideline Focused Update on Antithrombotic Therapy in Patients With Coronary Artery Disease. Circ J. 2020;84:831–865. doi: 10.1253/circj.CJ-19-1109

8. Gallone G, Baldetti L, Pagnesi M, Latib A, Colombo A, Libby P, Giannini F. Medical Therapy for Long-Term Prevention of Atherothrombosis Following an Acute Coronary Syndrome: JACC State-of-the-Art Review. J Am Coll Cardiol. 2018;72:2886–2903. doi: 10.1016/j.jacc.2018.09.052

9. Kulik A, Ruel M, Jneid H, Ferguson TB, Hiratzka LF, Ikonomidis JS, Lopez-Jimenez F, McNallan SM, Patel M, Roger VL, et al. Secondary prevention after coronary artery bypass graft surgery: a scientific statement from the American Heart Association. Circulation. 2015;131:927–964. doi: 10.1161/CIR.0000000000000182

10. Yasuda S, Kaikita K, Akao M, Ako J, Matoba T, Nakamura M, Miyauchi K, Hagiwara N, Kimura K, Hirayama A, et al. Antithrombotic Therapy for Atrial Fibrillation with Stable Coronary Disease. N Engl J Med. 2019;381:1103–1113. doi: 10.1056/NEJMoa1904143

11. Yasuda S, Kaikita K, Ogawa H, Akao M, Ako J, Matoba T, Nakamura M, Miyauchi K, Hagiwara N, Kimura K, et al. Atrial fibrillation and ischemic events with rivaroxaban in patients with stable coronary artery disease (AFIRE): Protocol for a multicenter, prospective, randomized, open-label, parallel group study. Int J Cardiol. 2018;265:108–112. doi: 10.1016/j.ijcard.2018.04.131

12. Vemulapalli S, Hellkamp AS, Jones WS, Piccini JP, Mahaffey KW, Becker RC, Hankey GJ, Berkowitz SD, Nessel CC, Breithardt G, et al. Blood pressure control and stroke or bleeding risk in anticoagulated patients with atrial fibrillation: Results from the ROCKET AF Trial. Am Heart J. 2016;178:74–84. doi: 10.1016/j.ahj.2016.05.001

13. Group SR, Wright JT, Jr., Williamson JD, Whelton PK, Snyder JK, Sink KM, Rocco MV, Reboussin DM, Rahman M, Oparil S, et al. A Randomized Trial of Intensive versus Standard Blood-Pressure Control. N Engl J Med. 2015;373:2103–2116. doi: 10.1056/NEJMoa1511939

14. Bi Y, Li M, Liu Y, Li T, Lu J, Duan P, Xu F, Dong Q, Wang A, Wang T, et al. Intensive Blood-Pressure Control in Patients with Type 2 Diabetes. N Engl J Med. 2025;392:1155–1167. doi: 10.1056/NEJMoa2412006

15. Kaikita K, Yasuda S, Akao M, Ako J, Matoba T, Nakamura M, Miyauchi K, Hagiwara N, Kimura K, Hirayama A, et al. Bleeding and Subsequent Cardiovascular Events and Death in Atrial Fibrillation With Stable Coronary Artery Disease: Insights From the AFIRE Trial. Circ Cardiovasc Interv. 2021;14:e010476. doi: 10.1161/CIRCINTERVENTIONS.120.010476

16. Pisters R, Lane DA, Nieuwlaat R, de Vos CB, Crijns HJ, Lip GY. A novel user-friendly score (HAS-BLED) to assess 1-year risk of major bleeding in patients with atrial fibrillation: the Euro Heart Survey. Chest. 2010;138:1093–1100. doi: 10.1378/chest.10-0134

17. Skanes AC, Healey JS, Cairns JA, Dorian P, Gillis AM, McMurtry MS, Mitchell LB, Verma A, Nattel S, Canadian Cardiovascular Society Atrial Fibrillation Guidelines C. Focused 2012 update of the Canadian Cardiovascular Society atrial fibrillation guidelines: recommendations for stroke prevention and rate/rhythm control. Can J Cardiol. 2012;28:125–136. doi: 10.1016/j.cjca.2012.01.021

18. Chhatriwalla AK, Amin AP, Kennedy KF, House JA, Cohen DJ, Rao SV, Messenger JC, Marso SP, National Cardiovascular Data R. Association between bleeding events and in-hospital mortality after percutaneous coronary intervention. JAMA. 2013;309:1022–1029. doi: 10.1001/jama.2013.1556

